# Is it time for a paradigm shift? Tailored online video education instead of pretest genetic counseling facilitates high genetic test uptake and informed choice for adults seeking cardiovascular genetic testing

**DOI:** 10.64898/2026.05.28.26354394

**Authors:** Bryana J. Rivers, Brittney Murray, Carolyn Applegate, Crystal Tichnell, Catherine Gordon, Rebecca McClellan, Emily E. Brown, Kathryn Nunez, Andreas S. Barth, Casey Overby Taylor, Lisa R. Yanek, Jeffrey Day, Cynthia A. James

**Author notes:** <u>Corresponding author</u>: Cynthia A. James, PhD, CGC Division of Cardiology, Department of Medicine, Johns Hopkins University School of Medicine 600 N. Wolfe St., Carnegie 568D Baltimore, MD, 21287.

## Abstract

**Background:** Pretest genetic counseling (GC) is recommended in conjunction with genetic testing (GT) for cardiovascular (CV) indications, yet access to CVGC is limited leading to delayed GT. Posttest GC could increase GC and GT access but requires efficient pretest education that supports both informed GT decision-making and robust GT uptake.

**Methods:** We developed four indication-tailored online CV genetics education videos and deployed them in a 3-arm randomized trial comparing pretest vs. posttest outpatient CVGC (RESEQUENCE-GC, NCT05422573). Participants were 1:1:1 randomized to pretest video education plus an optional (efficiency arm) or required (flipped arm) phone call with a genetic counselor and planned posttest CVGC or to standard pretest CVGC (SOC arm). Questionnaires administered at baseline and post-education included the CV Multidimensional Model of Informed Choice [MMIC] to quantify GT knowledge and informed GT choice.

**Results:** 389/767 (50.7%) adults aged 18-80 (mean 51.2±14.9 years) scheduling a first CVGC appointment consented to RESEQUENCE-GC and completed the baseline questionnaire. Efficiency arm participants (video education + optional phone call) were most likely to complete pretest education (134, 97.4% efficiency; 107, 85.6% flipped; 111, 87.4% SOC, p=0.0012) and elect GT (131, 95.6% efficiency; 105, 84.0% flipped; 107, 84.2% SOC, p=0.0036). Few (4, 2.9%) efficiency arm participants requested an optional pretest phone call. Most flipped arm participants (90, 84.1%) had no post-video questions, consistent with the 97 second [IQR: 65s-145s] median call duration. CV genetics knowledge was high post-education (median 8 [IQR 7,8]/8 MMIC items correct). Only video-based pretest education was associated with a significant increase in knowledge (p<0.0001). Nearly all participants made an informed GT choice with no difference between intervention (95.6%) and SOC (90.4%) arms (p=0.074).

**Conclusions:** Tailored, online video pretest education can enhance CV GT uptake, support informed GT decision-making, and be integrated into efficient pretest workflows, suggesting utility in scalable posttest CVGC.

## INTRODUCTION

Genetic counseling and testing for inherited cardiovascular conditions are recommended by professional organizations and have become increasingly more commonplace.^1–4^ As a means to narrow the differential and make appropriate management decisions, cardiologists commonly refer their patients for genetics services.^5^ Cardiovascular genetic counseling (CVGC) is guideline-recommended and associated with improved patient outcomes, however cardiologists report limited access to board-certified genetic counselors and increasingly longer wait times.^6–12^

The standard genetic counseling model likely contributes to these delays. Genetic counseling practice has traditionally utilized a pretest counseling model.^13,14^ During a pretest appointment a genetic counselor provides a standardized overview of the risks, benefits, limitations, and logistics of genetic testing.^15^ Genetic test results are then returned by phone and/or electronic medical record, or during a short posttest genetic counseling appointment.^11^ Alternatives to pretest one-to-one genetic counseling by a genetic counselor including webinars, chatbots, and videos have begun to be explored with the goal of reducing wait times and obtaining medically actionable genetic test results in a timely manner while still supporting informed genetic test decision-making.^11,16,17^

An informed choice is defined as a decision where: 1) knowledge is high and 2) the choice made is consistent with the person’s values or attitudes towards the issue at hand.^17–19^ Pretest education delivered by video has been shown to facilitate genetic test knowledge and high genetic test uptake in diverse oncology settings.^20–22^ In adult cardiology, video education has been implemented to promote informed choice or shared decision-making in other areas of cardiovascular care, including cardiac imaging, cardiac surgery, and invasive procedures (e.g. coronary angiography).^23–25^ It stands to reason that video education could facilitate informed choice for cardiovascular genetic testing as well. However, it is uncertain whether high-quality, tailored pretest genetics education videos can 1) support informed choice to a similar extent as pretest counseling by a genetic counselor, and 2) be integrated into efficient pretest genetics service workflows that support similar genetic test uptake.

To address these questions, herein we describe the development of pretest genetics education videos for four common cardiovascular genetic testing indications and evaluate their efficacy in supporting informed genetic test decision-making when deployed in the context of an ongoing three-arm randomized trial of pre- vs. posttest cardiovascular genetic counseling models (RESEQUENCE-GC, clinicaltrials.gov NCT05422573, registered 06/14/2022). In RESEQUENCE-GC educational videos plus an optional (efficiency arm) or required (flipped arm) phone call with a genetic counselor replace pretest genetic counseling in the intervention arms while the standard of care arm includes usual pretest genetic counseling by a genetic counselor. In this analysis we compare completion of pretest education, genetic test knowledge, informed choice, and genetic testing decision between the intervention and standard of care arms and describe the utilization, content, and duration of the optional/required pretest phone calls with a GC in the intervention arms.

## METHODS

This manuscript describes development and formative evaluation of cardiovascular genetic test education videos and their deployment in place of standard pretest genetic counseling by a genetic counselor in the context of a randomized trial. All elements of these studies have been approved by the Johns Hopkins Medicine IRB (IRB00320656, IRB00309500; PI James). Informed consent was provided by study participants. A portion of deidentified data can be made available upon the written request to the PI.

### Educational Video Development, Content, and Formative Evaluation

We developed four pretest genetics education videos tailored to both clinical indication (cardiomyopathy/arrhythmia [CM/Arr] or familial hypercholesterolemia [FH]) and genetic testing context (next generation sequencing panel testing or family-specific variant testing) using principles from the Cognitive Theory of Multimedia Learning.^26^ Written scripts were developed to include typical pretest genetic counseling content and subsequently refined by practicing cardiovascular genetic counselors. We collaborated with a medical illustrator (JD) to enhance the videos with illustrations and animations, which have been shown to boost engagement.^27^ The final videos featured both animations and scripted recordings of our center’s clinical cardiovascular genetic counselors explaining genetics concepts, genetic test utility, benefits and limitations, and genetic testing logistics (**Figure 1)**. Videos were created using Adobe Creative Cloud (Premiere, After Effects, and Illustrator).

**Figure 1.**
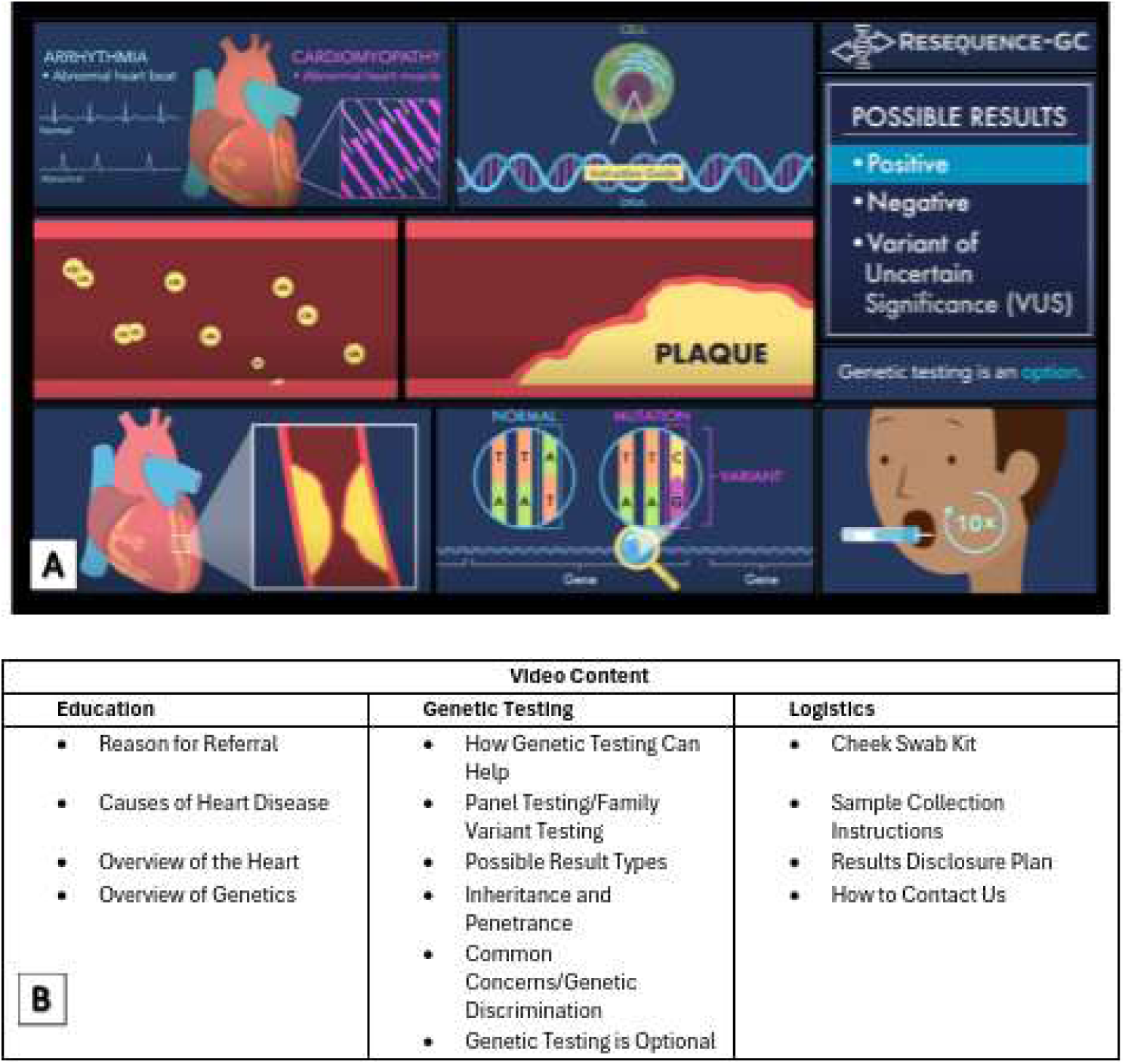
Genetics Education Video Content Panel A includes select screenshots from the cardiomyopathy/arrhythmia videos and the familial hypercholesterolemia videos. Panel B includes themes covered within the genetics education videos.

With the goal of replacing the education and informed consent portion of the pretest CVGC visit, it was important that the pretest videos address important concepts that patients should be aware of prior to proceeding with genetic testing that are included in a standard pretest visit. The Clinical Genome Resource (ClinGen) Consent and Disclosure Recommendations Workgroup (CADRe) outlines the following list of eight core concepts necessary for informed consent in clinical genetic testing: (1) genetic testing is voluntary; (2) the reason for referral and what is being tested; (3) the type of results and to whom they will be returned; (4) other potential results and available choices; (5) management implications; (6) potential impact on relatives; (7) limitations and next steps; and (8) potential risk of genetic discrimination and legal protections.^28^ The videos were aligned with these eight core concepts and they are each critical themes that we focus on in our standard clinical CVGC practice.

The videos were piloted and underwent iterative review by the RESEQUENCE-GC advisory board (clinicians, methods experts, and patients), members of the general public, and affected individuals recruited through advocacy groups (Hope for ARVD and Family Heart Foundation). Overall, the videos performed well in piloting, with high cardiovascular genetics knowledge as measured on the Cardiovascular or FH Multidimensional Model of Informed Choice knowledge scales and significant increases in knowledge after viewing a video^29–31^. Feedback led to refinements including shortening the videos, adding closed captions and end-of-video summaries, and adjusting vocal tone. Details of formative evaluation are provided in the **Supplemental Methods**. The final CM/Arr panel video was 12 minutes (m) and 32 seconds (sec) and the FH panel video was 12 m 25 sec. The final CM/Arr and FH family variant videos were 8m 18sec and 8m 47sec, respectively. Videos can be viewed at:

1. Cardiomyopathy Arrhythmia Panel Video
2. Familial Hypercholesterolemia Panel Video
3. Cardiomyopathy Arrhythmia Family Variant Video
4. Familial Hypercholesterolemia Family Variant Video.

### RESEQUENCE-GC Overview

The final videos were deployed for pretest education of participants in RESEQUENCE-GC (*Randomized clinical trial for the sequence of genetic counseling and testing to optimize efficiency, patient empowerment and engagement, and medical adherence for cardiac genetic testing indications*, NCT05422573). RESEQUENCE-GC is a three-arm parallel design randomized trial to evaluate the effectiveness of shifting the primary genetic counselor-patient interaction from pretest to posttest. In RESEQUENCE-GC intervention arms, pretest counseling with a genetic counselor is replaced with video education and either an optional (efficiency arm) or required (flipped arm) phone call with a genetic counselor before genetic testing. The primary CVGC appointment then occurs posttest when genetic test results were available. The third (standard of care) arm includes a pretest CVGC appointment with a genetic counselor. A CONSORT diagram detailing RESEQUENCE-GC design and trial enrollment is presented in **Supplementary Figure 1**. Enrollment, genetic counseling, and genetic testing have been completed for RESEQUENCE-GC. Enrollment began December 20, 2022 and ended August 15, 2025 with the final RESEQUENCE-GC-associated CVGC appointment in November 2025.

### Recruitment

Adults being scheduled for outpatient CVGC at the Johns Hopkins Center for Inherited Heart Diseases, located in a tertiary, urban, US academic medical center, were invited to participate in RESEQUENCE-GC based on the following eligibility criteria: 1) 18+ years, 2) English as a primary language, 3) no prior CVGC, 4) referred for CM/Arr or FH genetic testing including either panel testing or family-specific variant testing, 5) their upcoming CVGC appointment was at least four weeks away, and 6) their medical insurance did not require a pretest genetic counseling visit prior to genetic testing for coverage. Interested participants were further screened for eligibility by genetic counselor review of the electronic health record prior to contacting participants for oral consent. After providing oral consent for randomization, participants were 1:1:1 block randomized via a REDCap-based randomization list to one of three study arms stratified by panel vs. family specific variant testing with members of a family allocated to the same arm. Immediately following randomization, participants were sent a welcome email including a web link that directed them to their e-consent form (consenting to data collection) and online baseline questionnaire (Q1). Participants were offered a $200 Amazon® gift card for completing all relevant study questionnaires and were reassured a gift card would be provided regardless of genetic testing choice. All RESEQUENCE-GC participants who completed Q1 are included in these analyses.

### Pretest Education and Genetic Test Choice

For efficiency (optional phone call) and flipped (required phone call) arm participants, a clinical and genetic test indication-specific video launched immediately upon completion of their baseline questionnaire (Q1). After viewing their video, efficiency arm participants were asked to indicate whether they would like to proceed with genetic testing, and were presented with three options: “Yes,” “No,” or “I would like to talk to a genetic counselor first.” Flipped arm participants were prompted with the choice to proceed with the next steps or not. A “yes” response notified the study genetic counselor that the participant was awaiting their required phone call and flipped arm participants then elected whether to proceed with genetic testing during this phone call. The content and duration of the optional and required study phone calls were recorded. Standard of care participants attended a usual outpatient pretest CVGC appointment with one of our center’s four cardiovascular genetic counselors during which they decided whether to proceed with genetic testing. In all arms, after accepting or declining genetic testing, participants were invited to complete the post-education (Q2) questionnaire via email with an embedded link to the questionnaire. The email prompted patients to complete Q2 as soon as possible, prior to the receipt of their genetic test results. Four email reminders to complete Q2 were sent at 3-day intervals. Eligibility to complete Q2 ended upon return of genetic test results.

### Measures

#### Clinical and Demographic Data

Demographic data including age, race, ethnicity, gender, and highest level of education were self-reported by participants in the baseline questionnaire (Q1). Reason for referral, diagnosis, and type of genetic testing indicated, were obtained from genetic counselor review of participants’ cardiology records at the time of referral.

#### Genetic Test Education Process and Genetic Testing Decision

We recorded whether each participant completed pretest education. Completion of pretest education was defined as attending the pretest CVGC appointment in the standard of care arm, watching the video and completing the required phone call in the flipped arm, and watching the video and completing the optional phone call when requested in the efficiency arm. The point at which education was discontinued was recorded for each participant. Choice of whether to accept or decline genetic testing was recorded for each participant who completed pretest education.

#### Utilization, Content, and Duration of Pretest Calls with a Genetic Counselor (Intervention Arms)

We recorded whether each efficiency arm participant elected a pretest phone call. The content and duration of the optional (efficiency arm) and required (flipped arm) pretest phone calls with a genetic counselor were recorded both in participants’ electronic health records and in their study records. The content of the phone calls was categorized into themes and quantified. Examples include but are not limited to: no questions; clarifying question(s); testing logistics; and genetic testing and/or genetic counseling insurance coverage.

#### Genetic Test Knowledge and Informed Choice for Genetic Testing

Genetic test knowledge and attitudes were measured at Q1 and Q2 using the cardiac and FH Multidimensional Model of Informed Choice (MMIC) scales. The MMIC is a validated 13-item patient-reported outcome measure designed to assess patient knowledge and attitudes towards testing in a healthcare setting.^21^ It includes an 8-item true/false knowledge scale and a 5-item measure of attitudes on a 5-point (0-4) Likert scale. Each version of the MMIC utilizes a tailored indication-specific knowledge scale. We used version 1 of the recently validated “cardiac genetic testing” measure for participants with a CM/Arr genetic test indication as this version of the scale was created to be appropriate for individuals being offered genetic testing for potentially inherited arrhythmias, cardiomyopathies, and aortopathies.^31^ Different versions were used for probands (panel testing) vs. relatives (family specific variant testing) as the wording of two questions differs. For patients with a FH genetic testing indication, we used the FH MMIC knowledge scale which has a single battery of true/false questions. Validation of the FH MMIC scale was recently completed with publication pending (personal communication, Susan Christian, MSc, PhD). Attitudes and values questions are the same across MMIC scales.

Informed choice was derived from MMIC responses as has been done previously.^32,33^ An informed choice is considered one in which 1) knowledge is adequate and 2) the choice made aligns with preferences and values expressed. Knowledge was defined as answering ≥75% (6/8) of the true-false questions correctly as recommended by the cardiac genetic testing MMIC scale creators (S. Christian personal communication). Pre- and post-education genetic test knowledge was calculated based on the number of correct responses to the MMIC true/false questions in the Q1 and Q2 questionnaires respectively. Change in knowledge is the difference between the number of correct responses at Q1 and Q2. For attitude, we summed the values of each response to the attitude MMIC responses at Q2. Responses summing to ≥12 were considered a positive post-education attitude towards genetic testing. Finally, we categorized each participant by informed choice for genetic testing. Any participant with fewer than 6 correct answers on the T/F battery was considered to have made an uninformed choice because there was a lack of knowledge. In addition, participants whose knowledge score was ≥6 whose choice about genetic testing did not match their expressed preferences and values were considered to have made an uninformed choice. Informed choices in the setting of adequate knowledge included participants who 1) had a positive attitude towards genetic testing and decided to proceed with genetic testing or 2) had a negative attitude towards genetic testing and decided not to proceed with genetic testing.

### Statistical Analyses

Demographic and clinical characteristics were summarized using descriptive statistics. Completion of genetic test education and genetic test uptake were compared across study arm using Chi-square tests. The statistician (LY) and study PI were blinded to study group during analysis. Pre- and post-education knowledge scores for each MMIC knowledge subscale were calculated and compared via the Wilcoxon matched-pair signed-rank test. Comparison of change in knowledge in the standard of care vs. intervention arms was assessed using the Mann-Whitney U test. The proportion of participants making an informed choice for genetic testing was compared across study arms via Chi-square tests. Comparison of the distribution of clinical and demographic characteristics across study arms (e.g. adequacy of randomization) and their association with genetic test decision and informed choice were compared via Chi-square or Fisher’s exact tests as appropriate. Analyses were conducted in SAS (v9.4, SAS Institute Inc., Cary, NC) and SPSS (version 31; SPSS Inc., Chicago, IL). A p value <0.05 was considered statistically significant in all analyses. Primary prespecified RESEQUENCE-GC outcomes for which the trial was powered include psychosocial well-being and medical adherence 6 months post-test and will be reported separately.

## RESULTS

### Participants

Among 767 eligible patients, 389 (50.7%) consented to study participation and completed at least the baseline questionnaire (Q1). Consented participants were primarily non-Hispanic white (N=272, 69.9%) or Black (N=73, 18.8%), slightly more than half were female (N=224, 57.6%), and two-thirds had at least a bachelor’s degree (N=258, 66.3%) (**Table 1**). Participant age ranged from 18 to 80 with an average of 51.2±14.9 years. Most participants (N=269; 69.2%) were either probands (N=230) or family members (N=39) referred for genetic testing for a CM/Arr indication (**Table 1).** There were no significant differences in the distribution of demographic characteristics or clinical genetic testing indications among study arms (age: p=0.81; gender: p=0.10; race/ethnicity: p=0.37; education: p=0.47; clinical diagnosis: p=0.50; genetic test indication: p=0.08).

**Table 1.** Participant Demographics.

| Demographics (N = 389) |  | N (%) Total |
| --- | --- | --- |
| **Age** | 18-24 | 18 (4.6%) |
|  | 25-34 | 39 (10.0%) |
|  | 35-44 | 74 (19.0%) |
|  | 45-54 | 93 (23.9%) |
|  | 55-64 | 77 (19.8%) |
|  | 65-74 | 70 (18.0%) |
|  | 75-80 | 18 (4.6%) |
| **Gender Identity** | Male | 162 (41.6%) |
|  | Female | 224 (57.6%) |
|  | Non-binary/third gender | 1 (0.3%) |
|  | Prefer not to say | 2 (0.5%) |
| **Race** | Black/African American | 75 (19.3%) |
|  | White/European American | 281 (72.2%) |
|  | American Indian/Native American | 0 (0.0%) |
|  | Asian | 19 (4.9%) |
|  | Native Hawaiian/Pacific Islander | 2 (0.5%) |
|  | Mixed/Multi-Racial | 8 (2.1%) |
|  | Other/Not Listed | 4 (1.0%) |
| **Ethnicity** | Hispanic | 14 (3.6%) |
|  | Non-Hispanic | 375 (96.4%) |
| **Highest Level of Education** | Some high school | 3 (0.8%) |
|  | High school diploma | 25 (6.4%) |
|  | Trade, technical, or vocational training | 15 (3.9%) |
|  | Some college (no degree) | 62 (15.9%) |
|  | Associate's degree | 26 (6.7%) |
|  | Bachelor's degree | 114 (29.3%) |
|  | Any graduate degree | 144 (37.0%) |
| **Affected Proband** | **CM/Arr (N = 230)** |  |
|  | HCM | 94 (40.9%) |
|  | ACM | 13 (5.7%) |
|  | DCM | 71 (30.9%) |
|  | Channelopathy | 9 (3.9%) |
|  | Afib | 33 (14.3%) |
|  | Other CM/Arr | 10 (4.3%) |
|  | **Lipids (N = 117)** |  |
|  | FH/CAD | 117 (100%) |
| **Family Variant** | **CM/Arr (N = 39)** |  |
|  | HCM | 6 (15.4%) |
|  | ACM | 24 (61.5%) |
|  | DCM | 6 (15.4%) |
|  | Channelopathy | 3 (7.7%) |
|  | **Lipids (N = 3)** |  |
|  | FH | 3 (100%) |

Participant Demographics (N=389). HCM = Hypertrophic Cardiomyopathy; ACM = Arrhythmogenic Cardiomyopathy; DCM = Dilated Cardiomyopathy; Afib = Atrial Fibrillation; CM/Arr = Cardiomyopathy/Arrhythmia; FH = Familial Hypercholesterolemia; CAD = Coronary Artery Disease

### Genetic Test Education and Genetic Test Decision

While most participants (352, 90.5%) completed pretest education, there was a significant difference in likelihood across study arms (p=0.0012). As shown in **Figure 2**, 111 (87.4%) participants in the standard of care arm attended their pretest CVGC appointment. Similarly, 107 (85.6%) participants in the flipped arm completed pretest education. Among those who did not, 12/18 (66.7%) watched the genetic education video but did not complete their required phone call. In contrast, nearly all (N=134, 97.8%) participants in the efficiency arm completed pretest education, including watching the educational video and completing a phone call with a genetic counselor if requested.

**Figure 2.**
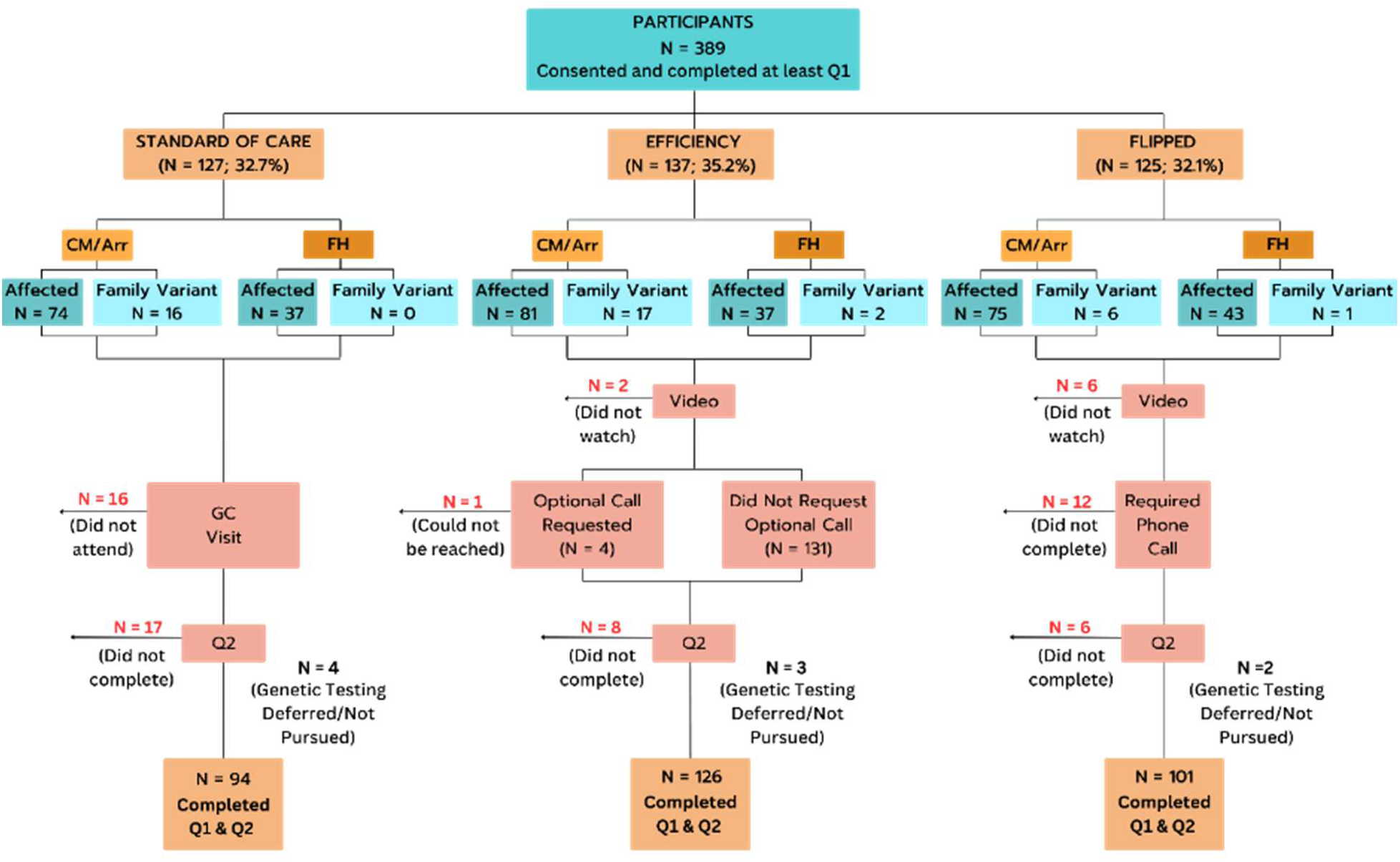
RESEQUENCE-GC pretest education and genetic test uptake 389 participants e-consented and completed at least Q1. Participants in each study arm were filtered out of analysis at various timepoints due to incompletion of study materials/processes. Ultimately 94 Standard of Care Arm, 126 Efficiency Arm, and 101 Flipped Arm participants completed both Q1 and Q2, for a total of 321 participants who completed both questionnaires.

Among the 352 participants who completed pretest education nearly all (N=343, 97.4%) elected to proceed with genetic testing with four (3.6%) in the standard of care arm, three (2.2%) in the efficiency arm, and two (1.9%) in the flipped arm declining or deferring genetic testing. Of note, one of the two participants in the flipped arm who did not initially proceed with genetic testing later recontacted the genetics team to pursue testing after securing life insurance. Ultimately, participants in the efficiency arm had a significantly higher genetic test uptake with 131 (95.6%) choosing to proceed with genetic testing compared to 107 (84.2%) in the standard of care and 105 (84.0%) in the flipped arms (p=0.0036).

#### Flipped Arm Phone Calls (Required)

Most participants in the flipped arm who completed their required phone call with the study genetic counselor after watching the genetic test education video had no questions (N=90/107, 84.1%). Reflecting this, the median call length was 97 seconds [IQR: 65s-145s]. Two additional participants (1.9%) did not have specific questions but acknowledged that the video prompted them to postpone genetic testing due to life insurance concerns. Most questions asked did not reflect a lack of understanding of the content of the videos. Four participants had basic clarifying questions; four had testing logistics questions; four participants wondered about insurance coverage of the genetic testing and/or genetic counseling visit; one participant wanted to confirm their study eligibility; and only two participants required reiteration of the contents of the video. Topics of clarifying questions included: if a mutation could develop later; laboratory sample retention policies; testing logistics and considerations for relatives; if testing could determine if a variant is maternally or paternally inherited; and how much of a person’s DNA is passed onto their children. Content that required reiteration included study process flow and purpose of the genetic testing. The frequency and content of questions are summarized in **Figure 3**.

**Figure 3.**
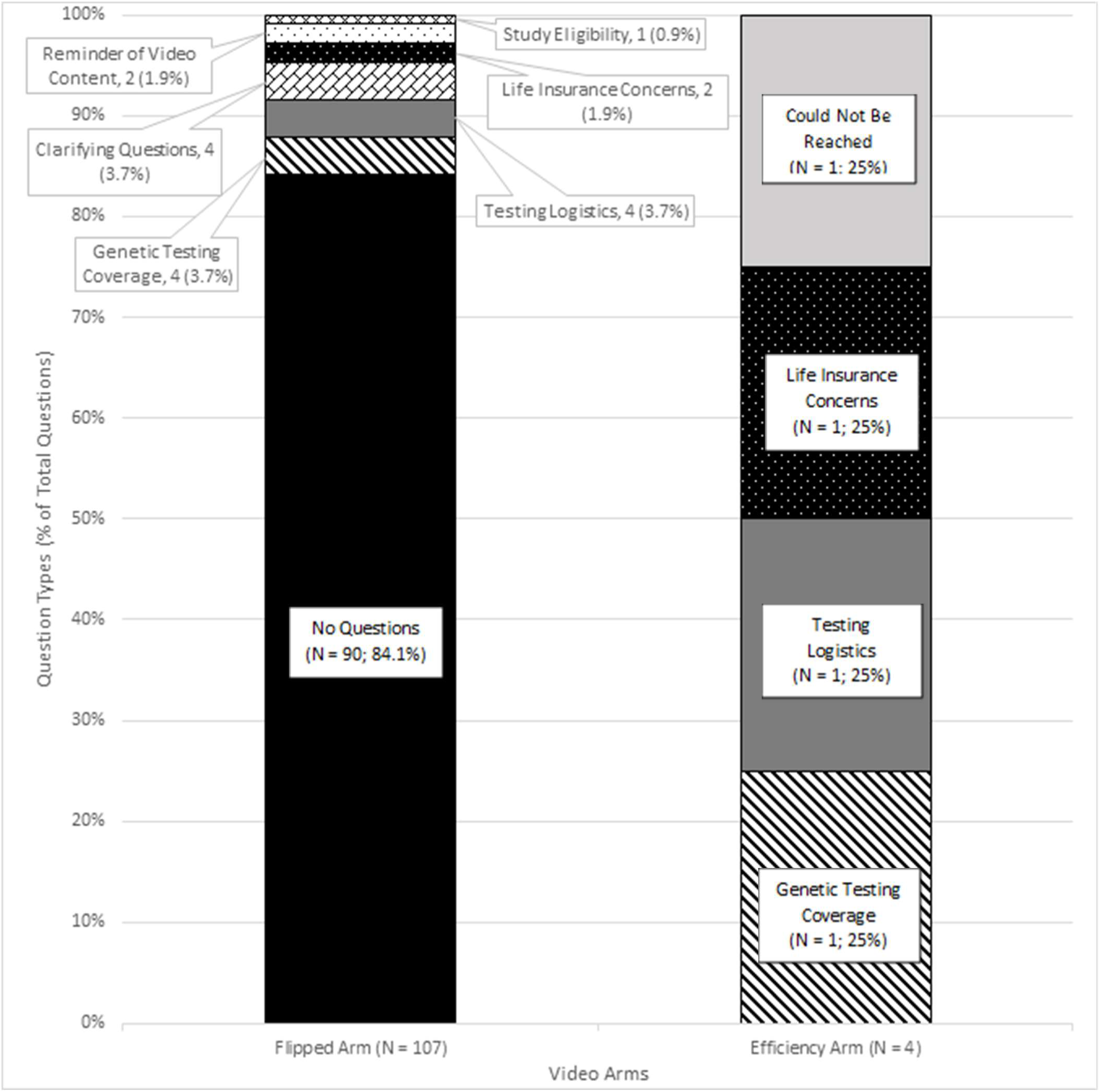
Content of Required and Optional Phone Calls This chart summarizes the content of the questions that participants asked the genetic counselor during their required (flipped arm) or optional (efficiency arm) phone calls. The number and percentage of question types are reflected in the stacked bar charts.

#### Efficiency Arm Phone Calls (Optional)

Few efficiency arm participants (N=4, 2.9%) requested a phone call with a genetic counselor prior to deciding about genetic testing (**Figure 3**). One requested the call to indicate that the video facilitated their decision *not* to proceed with genetic testing (e.g. the video prompted life insurance considerations), a second participant had very case-specific testing logistics inquiries and later canceled their CVGC visit due to insurance coverage, and a third had questions about genetic testing coverage. One final participant could not be reached for their requested phone call after several attempts but later emailed the study team asking further questions about health and life insurability after watching the video.

### Knowledge and Informed Choice

Of the 352 participants who completed pre-test education, 321 (91.2%) completed the post-education questionnaire (Q2) and were therefore included in analyses of genetic test knowledge and informed choice. As shown in **Supplementary Table 1**, participants were knowledgeable about cardiovascular genetic testing and the number of correct responses on the MMIC knowledge scale increased significantly following pretest education (p=0.00027). While there were significant increases in the number of correct answers among participants who had video education (p=0.00033), there was no significant change for patients who received standard of care pretest CVGC (p=0.27). Notably, two-thirds of participants (N=214, 66.7%) answered all eight of their MMIC questions correctly post-education, a significant increase from the proportion doing so pre-education (p<0.0001, McNemar). Consistent with these findings, nearly all participants (N=314, 97.8%) had a post-education MMIC knowledge score ≥ 6, (≥75% correct) and thus fulfilled the criteria for being sufficiently knowledgeable about key principles of genetic testing to have the capacity to make an informed choice (**Table 2**).

**Table 2.** Knowledge and Informed Choice.

| MMIC post-education<br>(Score ≥75%) | Frequency<br>(N = 321) | Attitudes and Testing Decision |  |  |
| --- | --- | --- | --- | --- |
| Yes | 314 (97.8%) |  | Genetic Testing Accepted? |  |
|  |  | Positive Attitude About Genetic Testing? | Yes | No |
|  |  | Yes | 301* | 3<sup>†</sup> |
|  |  | No | 9<sup>†</sup> | 1* |
| No | 7<sup>†</sup> (2.2%) | --- |  |  |
| * = Informed Choice; † = Uninformed Choice |  |  |  |  |
| Informed Choice | Frequency<br>(N = 321) | Study Arm |  |  |
|  |  | Standard of Care<br>(N = 94) | Intervention<br>Arms<br>(N = 227) | P-Value |
| Yes | 302 (94.1%) | 85 (90.4%) | 217 (95.6%) | 0.074 |
| No | 19 (5.9%) | 9 (9.6%) | 10 (4.4%) | -- |

Abbreviations: MMIC: Multidimensional Measure of Informed Choice

Furthermore, as shown in **Table 2**, nearly all (N=302, 94.1%) knowledgeable participants demonstrated informed choice by making a genetic testing decision that was consistent with their attitudes about genetic testing. Overall, informed choice was high across all study arms, with 85 (90.4%) standard of care, 95 (94.1%) flipped, and 126 (96.8%) efficiency arm participants classified as having made an informed choice regarding genetic testing. There was no significant difference in the likelihood of participants who had standard of care pretest CVGC (90.4%) vs. pretest education by video (95.6%) making an informed choice (p=0.074). Additionally, as shown in **Supplementary Table 2**, there was no association between sociodemographic characteristics or genetic testing indication and likelihood of making an informed choice.

## DISCUSSION

Genetic counseling for cardiovascular indications has become a guideline-recommended service as cardiovascular genetic testing becomes more commonplace.^1^ Cardiologists rely on collaboration with genetic counselors for appropriate genetic test selection, obtaining informed consent, and interpreting genetic test results.^12^ At present, access to genetic testing can be delayed by limitations in the availability of CVGC services and resultant long wait times.^34^ Genetic counselors and cardiologists alike recognize the value of improving access to care.^12,35^ One opportunity to do so is to increase efficiency by moving away from standard pretest genetic counseling by a genetic counselor, an approach which has been explored in other adult genetics sub-specialties.^22,36^ In cardiology, while prior studies have evaluated genetic counseling outcomes including patient empowerment, patient satisfaction, cardiac anxiety, genetic test uptake, and cascade screening for relatives, these observational studies have primarily utilized standard pretest CVGC and none have tested CVGC outcomes in a randomized trial.^6,37,38^

In the current randomized trial, we evaluated pretest video education and compared it to the standard of care model of CVGC wherein the bulk of genetic counselor time is spent during the pretest visit. We found that pretest video education with planned posttest results-focused CVGC did not negatively impact participant knowledge or attitudes about genetic testing compared to the standard of care model. In fact, we observed greater gains in knowledge following video education and found a high proportion of patients made an informed choice in all study arms with no significant difference in the video arms compared to the standard of care arm (**Table 2**). Furthermore, patients offered video-based education had a higher uptake of guideline-recommended genetic testing. In summary, participants were able to make an informed choice about cardiovascular genetic testing without a pretest genetic counseling appointment and without sacrificing, and even potentially improving, genetic test uptake.

The study was strengthened by the recent development and validation of the cardiac specific MMIC scales which allowed our study to more rigorously measure patients’ pre- and post-education cardiovascular genetics knowledge, which has historically been area of genetic counseling literature that presents a gap, particularly for CVGC.^31^ Informed consent is critical to pretest genetic counseling and informed choice is a widely-recognized pretest genetic counseling outcome. Therefore to comprehensively demonstrate the effectiveness of the cardiovascular genetics videos, patients’ ability to make an informed choice post-video was paramount to videos’ success as a pretest tool.^39^ Together, our results suggest that pre-test video education can promote high levels of important genetic test knowledge, informed choice for genetic testing, and, at least in the efficiency arm, high uptake of what are increasingly guideline-recommended genetic tests.

Our study included an optional or required post-video education phone call with a genetic counselor prior to genetic testing. While this may aid informed decision making for some patients, requiring a phone call is unlikely to be necessary in practice. As we observed with this study, there was a very limited uptake of optional phone calls, most participants had no questions during required phone calls, and the duration of required calls was very short. In fact, requiring the phone call was a rate-limiting step in the flipped arm as 14.4% of participants did not complete pretest education, mostly because they did not complete the required phone call. We do feel, however, that it is important for genetic counselors to remain available for *optional* post-video phone calls or visits to answer patients’ questions about health and life insurance, testing logistics, or other concerns that may arise. Genetic counselor expertise in providing tailored education and facilitating decision-making is an important component of CVGC that should remain available in a posttest model for patients who need it.^31^

### Limitations

By incorporating this study into our existing clinic structure, we were able to observe the benefits and pitfalls of a posttest genetic counseling model, as well as its potential real-life applicability. We had the benefit of a diverse population across race, age, and level of education in which to test these service delivery models (**Table 1**). It is important to note that overall, participant knowledge was already high at baseline. When implementing video education in a cardiovascular genetics clinic outside of the context of the study, all patients should be presented with the option for either pretest or posttest genetic counseling, but provider discretion may be necessary. In standard clinical practice, not all patients may be motivated to learn by video. Such individuals are likely under-represented in this study as they may have been more likely to decline participation in a study that uses video education. Additionally, the videos were created in the English language, so we were unable to assess the utility of pretest video education in other languages. The broader applicability of these videos beyond a single tertiary care center has not been assessed. While a participation incentive was necessary given the participant effort, it is not measurable what impact this may have had on genetic testing completion rates. Finally, this manuscript establishes that pretest video education leads to improved genetic test uptake with similar levels of knowledge and informed choice, which is necessary, but not sufficient for the success of posttest CVGC. Further RESEQUENCE-GC analyses will evaluate longer-term patient-reported psychosocial impact of genetic counseling and testing in a post-test model as well as longer term impact on medical adherence.

### Conclusions

A posttest model of genetic counseling has the potential to increase patient access to cardiovascular genetic testing and the efficiency of the CVGC and genetic testing process. These data show that genetic counselor-designed pretest education videos with a genetic counselor available to address questions supports pretest knowledge, facilitates informed choice, and increases genetic test uptake when used in combination with planned posttest genetic counseling. In turn, this may potentially help streamline the cardiovascular genetic testing process, allowing patients and their cardiovascular providers faster access to actionable genetic test results.

## Data Availability

A portion of deidentified data can be made available upon the written request to the PI.

## Acknowledgments

The authors would like to thank the RESEQUENCE-GC Advisory Board, the Family Heart Foundation, Hope for ARVD, and the patients who participated in this study.

## Supplemental Material

Supplemental Methods – Pre-test Education Video Development and Formative Evaluation Supplemental Tables and Figures (Tables S1–S2; Figure S1)

References 40-41

## Non-Standard Abbreviations and Acronyms

RESEQUENCE-GC: Randomized clinical trial for the sequence of genetic counseling and testing to optimize efficiency, patient empowerment and engagement, and medical adherence for cardiac genetic testing indications
CVGC: Cardiovascular Genetic Counseling
MMIC: Multidimensional Model of Informed Choice
CM/Arr: Cardiomyopathy/Arrhythmia
Q1: Baseline Questionnaire
Q2: Post-Education Questionnaire

